# Regional Anesthesia use in Pediatric Burn Surgery: A Retrospective Observational Cohort Study

**DOI:** 10.1101/2020.11.25.20231407

**Authors:** M Richman, J Berman, EM Ross

## Abstract

Approximately 77,000 children 16 years or less suffered burn injuries in the United States in 2018. Treatment, reconstruction, and rehabilitation are painful experiences. For some, the experience triggers post-traumatic stress disorder (PTSD) and/or a chronic pain syndrome. Given the role pain plays as a major secondary disease, it must be addressed to achieve optimal healing. Regional anesthesia has been used extensively to manage postoperative pain and reduce the need for opioids following other surgical procedures in children. Nevertheless, regional anesthesia has not yet been widely used in pediatric burn care. We present a demonstration project utilizing regional anesthesia in 15 pediatric burn patients over an eight-month period. Our results indicate that the use of regional anesthesia reliably reduces perioperative pain and opioid requirements in the immediate peri-operative period. In this cohort, 93% of patients scored a 0/10 on a FLACC scale for pain by post-anesthesia care unit (PACU) discharge, with an average PACU stay of 70 minutes. Thirty-three percent of patients received no opioids, and the average opioid dose was only 0.06mg/kg morphine equivalents. We conclude that regional anesthesia can be used to improve patient comfort and decrease opioid requirement.

## Introduction

Burns are defined as an injury or damage to the skin or other organic tissue resulting from exposure to thermal, chemical, radiologic, electrical or friction trauma.^1^ Burn injuries are the most common injury to pediatric patients in the world, accounting for approximately 2500 fatalities annually in the United States alone. Of those reported, scald burns are the main culprit in children younger than five years of age, while flame-related burns are more commonly associated with older children and adolescents.^2^ Burns are traumatic both physically and emotionally, especially to vulnerable children. Serial dressing changes, as well as physical and occupational therapy, cause repetitive trauma, particularly when pain is not well controlled.^3^ Poorly controlled pain has been shown to decrease compliance with treatment regimens; in addition pain can delay and degrade wound healing through altered neurohormonal responses. Wollgarten-Hadamek demonstrated that children aged nine to sixteen years who had suffered moderate to severe burns at age six to twenty-four months had long-term alterations in somatosensory and pain processing.^4^

Burn pain emanates through multiple pathways, and optimal pain control is often not achievable through opioid therapy alone. Regional anesthetic techniques can greatly decrease the pain levels experienced during burn debridement and dressing changes. Shank, et al found that children undergoing skin graft harvests who received nerve blocks at the donor site had less postoperative pain than those receiving surgical infiltration of local anesthetics.^5^ In addition to analgesia, the sympathectomy induced vasodilation of local anesthetics increases tissue blood flow and thus possibly improves local wound healing. Though little literature exists specific to burn healing, it has been shown that regional anesthesia decreases failure rate of arteriovenous fistulas via a similar mechanism.^6^ Further, it is well documented that regional anesthesia reduces opioid use.^7^ The benefits of this decreased opioid use are multifold: improved bowel function, decreased potential for short and long term opioid sensitization, decreased need for weaning and dependence, and reduced potential for long-term psychological effects.

This demonstration project establishes proof of concept that regional anesthesia can be successfully used to manage perioperative pain and reduce opioid utilization in pediatric burn care.

## Methods

We report a retrospective observational cohort study of pediatric burn patients who received regional anesthesia in conjunction with general anesthesia for surgical treatment of their burn injuries. We have no interests to declare. The IRB determined that this study did not require consent other than that for the anesthetics performed. The study occurred at a University tertiary health care system that is an American Burn Association verified burn center. Records of all pediatric burn patients treated over an eight-month period who required operative repair and received a regional anesthetic block were examined. Fifteen patients were identified. An analysis of the 15 patients was undertaken to determine regional block, intra-operative analgesic and anesthetic usage, as well as immediate post-operative analgesic usage, pain levels and time spent in the PACU.

## Results

Over an eight-month period, fifteen patients aged seven months to fourteen years received general anesthesia and an appropriate regional block based on location of burn and graft for burn debridement and grafting. All patients except one received an appropriate weight-based dose of Bupivacaine 0.25% with or without clonidine; the other received an appropriate weight-based dose of 0.5% Ropivacaine. All patients except one received approximately half minimum alveolar concentration (MAC) of sevoflurane for the duration of their anesthetic. Average intraoperative fentanyl dosing was 1.2 mcg/kg, with most patients receiving 0.5-0.9 mcg/kg. In addition, 9/15 patients received dexmedetomidine (average dose 0.6 mcg/kg) and 3/15 ketamine (average dose 0.75 mg/kg). Four patients received acetaminophen at either 10 mg/kg or 15 mg/kg based on their age. Two patients received 0.5 mg/kg of ketorolac. One-third of patients received no analgesic adjuvants in addition to the regional block and opioid.

When examining pain scores and PACU stay we found the following. Sixty-seven percent of patients had FLACC scores of 0 upon arrival to the PACU. This rose to 93% by discharge, with an average discharge time of 70 minutes (range 33-158 minutes). Average opioid use in the PACU ranged from 0-0.2mg/kg morphine equivalents with a mean of 0.06mg/kg. One-third of patients received no opioids in the PACU. Table 1 shows perioperative analgesic use, OR and PACU times as well as FLACC scores based on age. Table 2 stratifies these criteria in terms of block(s) used.

**Table 1:** Perioperative analgesic usage, pain scores and PACU times stratified by age

| Age Range | Number of Patients | Intraoperative Fentanyl Range (avg) mcg/kg | PACU Time Range (avg) Minutes | PACU | FLACC Range (avg) | Scores | PACU Morphine Equivalent Range (avg) mg/kg |
| --- | --- | --- | --- | --- | --- | --- | --- |
|  |  |  |  | 0 minutes | 15 minutes | Discharge |  |
| 7 to 13 months | 3 | 0-1.9 (0.6) | 33-69 (53) | 0 | 0-5 (2) | 0 | 0-0.03 (0.01) |
| 2 to 4 years | 4 | 0.5-0.9 (0.8) | 44-53 (48) | 0 | 0-8 (4) | 0 | 0-0.1 (0.03) |
| 5 to 8 years | 4 | 0.5-2.8 (1.8) | 44-158 (90) | 0-10 (5) | 0-6 (1.5) | 0 | 0.1-0.2 (0.1) |
| 11 to 14 years | 4 | 0.5-1.7 (1.2) | 55-100 (79) | 0-7 (2.5) | 0-9 (5) | 0-5 (1) | 0.04-0.15 (0.09) |

**Table 2:** PACU pain scores and analgesic usage stratified by block type *One patient received both supraclavicular and caudal blocks **One patient received both femoral and popliteal blocks

| Block | Number of Patients | Anesthesia Time Range (Avg) Minutes | PACU Time Minutes | PACU | FLACC | Scores | PACU Morphine Equivalent mg/kg |
| --- | --- | --- | --- | --- | --- | --- | --- |
|  |  |  |  | 0 minutes | 15 minutes | Discharge |  |
| *Supraclavicular | 4 | 74-129 (101) | 33-69 (51) | 0 | 0-5 (1) | 0 | 0-0.03 (0.01) |
| Transversus Abdominus Plane | 2 | 53-197 (125) | 46-100 (73) | 0-7 (4) | 0-4 (2) | 0-5 (2.5) | 0-0.09 (0.05) |
| **Femoral | 4 | 61-165 (106) | 46-158 (96) | 0-8 (4) | 0-6 (4) | 0 | 0-0.16 (0.08) |
| **Popliteal | 2 | 77-132 (105) | 46-75 (61) | 0 | 0 | 0 | 0.1-0.15 (0.13) |
| *Caudal | 2 | 116-124 (120) | 53-58 (56) | 0 | 0-8 (4) | 0 | 0-0.03 (0.02) |
| Lumbar epidural | 3 | 127-291 (216) | 44-114 (71) | 0-10 (4) | 0-9 (6) | 0 | 0.1-0.2 (0.12) |

## Discussion

Pediatric burn pain is difficult to control and often leads to long-term suffering. Undertreated acute pain, particularly in pediatric burn patients, can lead to chronic pain and psychiatric maladies. Multiple previous studies examining other surgeries have demonstrated the benefits of regional anesthesia in the pediatric population with regards to pain control and decreased anesthetic usage.^8^ In addition, patients have been shown to experience decreased urinary retention and post-operative nausea/vomiting as well as quicker discharge, return of ambulation, and overall recovery.^9^ Increased use of peri-operative opioids has been shown to lead to greater continued use.^10^ Additionally, concern has been raised that use of narcotic medications may potentially lead to adverse cognitive and psychomotor effects given the many opioid receptors in brain regions which moderate or modulate attention, memory, and learning tasks.^11^ It can be concluded that decreased peri-operative opioid utilization would prevent the physical and social consequences of abuse. Little data exists on regional anesthetic block use in pediatric burn injury patients. Our study demonstrated that the use of regional anesthesia can greatly reduce acute pain and anesthetic usage with little to no opioid use in the perioperative setting. In addition, patients were able to receive less anesthesia, which reduces hemodynamic fluctuations from sympathetic activation.

One major benefit highlighted in studies on regional anesthetic block use is decreased PACU length of stay.^8^ Shorter PACU times decrease hospital costs and improve operating room flow. While this retrospective series did not compare patient populations, our study population did experience objectively short PACU times, leading to an obvious benefit for the hospital as well as for the patient.

While this study focused on the immediate perioperative period, burn injury has multiple points of insult that include the initial burn as well as subsequent dressing changes. In addition to the initial injury, repetitive insults from treatments and dressing changes further traumatize children. Indeed, many children view dressing changes as the most traumatizing aspects of their burn injury.^12^ Children lack the ability to understand the long-term benefits of painful procedures and treatments; consequently, they are more susceptible to ensuant psychological maladies such as anxiety, depression, and post-traumatic stress disorder.^9^ Though preventing pain in the immediate post-operative period may decrease these outcomes,^13^ prolonging this effect would be even more beneficial. Local anesthetic regional blocks often lend themselves to catheter placement which would lengthen the benefits seen in this series. Further studies should expand upon this to improve outcomes in this patient population.

## Data Availability

Data was available from the University of North Carolina at Chapel Hill Hospital's Pediatric Pain and Regional Service patient archives. These are HIPPA protected locked charts and are not available to the public.

